# Leveraging Large Language Models for Colorectal Cancer Symptom Extraction from MIMIC-IV Clinical Notes

**DOI:** 10.64898/2026.09.15.26362961

**Authors:** Youran Lee, Ivo Dinov, Xiaosu Hu, Yun Jiang

## Abstract

**Background:** Much of the symptom burden in colorectal cancer (CRC) patients is documented in unstructured discharge-note narrative, and manual extraction is not scalable. Whether large language models (LLMs) outperform rule-based and named entity recognition (NER) methods has not been rigorously benchmarked.

**Objective:** To benchmark rule-based, NER, and zero-shot LLM methods for extracting 46 cancer-related symptoms from CRC discharge notes against an adjudicated ground truth.

**Methods:** We analyzed 2,704 discharge notes from CRC patients in MIMIC-IV. A 46-symptom target list was built from the Memorial Symptom Assessment Scale and the EORTC QLQ-CR29. Four approaches -- dictionary-based rule matching, pretrained clinical NER, and zero-shot Claude Haiku and Gemini 3.5 Flash -- plus two hybrid variants (LLM output with post-hoc rule-based negation filtering) were evaluated against a 200-note gold standard adjudicated by two raters (pooled kappa=0.71, macro kappa=0.49), using Macro/Micro F1, precision, and recall.

**Results:** Gemini 3.5 Flash performed best (Macro F1=0.70, Micro F1=0.86, Macro Precision=0.74), followed by Claude Haiku (Macro F1=0.63, Macro Recall=0.71); both substantially outperformed rule-based (Macro F1=0.44) and NER (Macro F1=0.38) methods. Post-hoc negation filtering paradoxically degraded LLM performance (Gemini+Hybrid Macro F1=0.58; Claude+Hybrid Macro F1=0.54) by overriding correct predictions through rigid, fixed-window matching.

**Conclusions:** Zero-shot LLMs substantially outperform rule-based and NER approaches for CRC symptom extraction; post-hoc negation correction should not be applied to LLM outputs without syntactic scope validation. Implications for Practice: Zero-shot LLM extraction offers a scalable, accurate alternative to manual chart review and traditional NLP pipelines for oncology symptom surveillance, without institution-specific rule development or model training.

## Introduction

Colorectal cancer (CRC) is the third most common cancer diagnosed in the United States.[1] Patients with CRC experience a range of disease- and treatment-related symptoms, including fatigue, rectal pain, feelings of incomplete bowel evacuation, constipation, and diarrhea, which can contribute to substantial symptom burden and adversely affect quality of life and clinical outcomes.[2, 3] Information about these symptoms is richly documented in electronic health records (EHRs) throughout routine clinical care. However, much of this information is embedded in unstructured narrative text authored by multidisciplinary providers, [4, 5] making it difficult to systematically access and analyze without advanced computational tools. Manual chart review is resource-intensive and difficult to scale beyond small samples. [6] Consequently, traditional symptom research has relied primarily on structured patient-reported outcome instruments or small chart-review studies. However, neither approach can systematically quantify how a broad, predefined set of symptoms is documented in narrative notes across large, real-world clinical populations.[7]

Natural language processing (NLP) offers one approach to unlocking this unstructured clinical narrative. Rule-based NLP (dictionary or pattern matching against curated term lists) and named entity recognition (NER) methods (which use a trained model to automatically tag clinically relevant spans of text) have both demonstrated feasibility for extracting symptom information from clinical text, but their performance is limited by sensitivity to variation in clinical language, negation, and contextual ambiguity. Large language models (LLMs) have recently emerged as promising tools for interpreting free-text clinical narrative, offering greater flexibility than rule-based or NER systems in handling varied phrasing, synonyms, and implicit symptom descriptions without requiring institution-specific rule development or model retraining.[8] Using zero- or few-shot prompting -- which requires no task-specific model training -- LLM-based methods have been applied across healthcare domains to identify clinical risk factors, extract diagnoses, and predict patient outcomes, including symptom extraction from free text in diverse patient populations, among them patients with cancer.[9, 10] However, few studies have directly benchmarked rule-based, NER, and LLM-based extraction against one another within the same patient population and the same expert-adjudicated ground truth, and rigorous multi-method benchmarking specific to a defined, clinically comprehensive symptom inventory remains limited.

To address this gap, the objective of this study was to develop and systematically benchmark rule-based, NER, and zero-shot LLM methods -- that is, to compare their extraction performance against a common, expert-adjudicated ground truth using standardized metrics -- for automated extraction of CRC-related symptoms from discharge notes of CRC patients in MIMIC-IV, a large, publicly available, de-identified hospital EHR database.

## Methods

### Data Source

This study utilized the MIMIC-IV Clinical Notes dataset (version 2.2), a large-scale de-identified clinical note database from Beth Israel Deaconess Medical Center.[11] Discharge summaries were extracted for patients diagnosed with colorectal cancer (CRC), identified using ICD-9 codes (153.x, 154.x) and ICD-10 codes (C18, C19, C20) from the MIMIC-IV hospital module. A total of 1,507 patients with colorectal cancer, and 2,728 discharge notes were included in the analysis.

### Symptom Measures

The target symptom list was constructed by integrating two validated patient-reported outcome instruments, selected to jointly provide both broad, cross-cancer symptom coverage and colorectal cancer-specific symptom coverage: the Memorial Symptom Assessment Scale (MSAS),[12] a widely used, multidimensional instrument capturing general physical and psychological cancer symptoms, and the European Organization for Research and Treatment of Cancer Quality of Life Questionnaire – CRC module-29 (EORTC QLQ-CR29),[13] a colorectal cancer-specific quality-of-life questionnaire capturing bowel, genitourinary, and stoma-related symptoms not well represented in generic cancer symptom instruments. After removing duplicates, the combined inventory yielded 46 unique symptoms spanning physical, psychological, and gastrointestinal domains.

### Text Preprocessing

For each discharge note, the Chief Complaint (CC) and History of Present Illness (HPI) sections were extracted using regular expressions that matched standard section-header strings (e.g., “Chief Complaint:,” “History of Present Illness:”) and captured the text following each header up to the next recognized section header. These sections were selected because they narratively summarize the patient’s current presentation and are where symptom-relevant information is most consistently documented, in contrast to other discharge-note sections (e.g., past medical history, discharge medications) that primarily document historical or administrative information. Notes for which CC/HPI extraction failed due to non-standard formatting or absent section headers (n=24) were excluded, resulting in 2,704 notes used in the primary analysis.

### Symptom Extraction Methods

Four primary approaches -- a dictionary-based rule-based method, a pretrained biomedical NER model,[14] and two zero-shot LLMs (Claude Haiku and Gemini 3.5 Flash) -- and two hybrid variants (LLM output with post-hoc rule-based negation filtering) were evaluated for automated symptom extraction. These approaches were selected to span the methodological spectrum used in clinical NLP, from fully transparent dictionary matching, through pretrained biomedical named entity recognition, to general-purpose LLMs that require no task-specific training, allowing direct comparison of their relative performance on the same extraction task and the same ground truth. All preprocessing, extraction pipelines, and evaluation were implemented in Python (version 3.10) using the following libraries: re (regular expressions) and pandas for the rule-based method; spaCy, scispaCy,[15] and negspaCy for the NER method; anthropic and google-generative AI for LLM API calls; and pandas and scikit-learn for data handling and evaluation metrics.

*Rule-based method.* A dictionary-based approach was developed: each of the 46 symptoms was matched against the discharge-note text using a curated list of clinical synonyms and abbreviations for that symptom (e.g., “dyspnea,” “SOB,” “shortness of breath,” and “difficulty breathing” for shortness_of_breath), derived from the same synonym/trigger-term list developed for the annotation coding manual (see Ground Truth Annotation; full synonym list and representative examples of how each symptom is documented in the notes are provided in Supplementary Table 1). Following the general approach of classic window-based clinical negation-detection algorithms,[16] a fixed list of 21 negation cues (Table 1) -- grouped into general negations, denial terms, absence/finding-negation phrases, resolution statements, and diagnostic-exclusion phrases -- was searched within a 100-character window preceding each matched symptom term; a match with a negation cue in this preceding window caused the mention to be excluded.

**Table 1.** Negation Cues Used in the Rule-Based Extraction Method.

| Category | Negation Cues | Position |
| --- | --- | --- |
| **General negations** | no, not, never, none, without, w/o | Before |
| **Denial terms** | denies, denied, deny | Before |
| **Absence / negative findings** | negative for, no evidence of, absent, absence of, free of, no signs of, no sign of, no complaints of, unremarkable for | Before |
| **Resolution statements** | resolved | Before |
| **Diagnostic exclusion** | ruled out, r/o | Before |

*Note.* All cues were matched as whole words/phrases within a 100-character window preceding the matched symptom term (Position = Before); the rule-based method did not use post-term negation cues.

*Named Entity Recognition (NER).* A pre-trained biomedical named entity recognition model (en_ner_bc5cdr_md), built on the spaCy NLP framework via the scispaCy library,[15] was applied to identify disease/problem entities -- spans of text corresponding to diseases, signs, and symptoms -- in the CC/HPI text. The model was trained on the BioCreative V Chemical-Disease Relation (BC5CDR) corpus,[17] a general biomedical entity-recognition dataset rather than a symptom-specific corpus. Detected entities were subsequently mapped to the study’s 46-symptom inventory during post-processing. Negation of each detected entity was determined using the negspaCy implementation of the ConText algorithm,[18] restricted to disease-type entities; negated entities were excluded from further processing. Non-negated entity text was matched against the same 46-symptom synonym dictionary used by the rule-based method (whole-word/phrase matching) to map entities onto the predefined 46-symptom inventory.

*Claude Haiku (LLM).* Claude Haiku (claude-haiku4.5, Anthropic18) was applied in a zero-shot setting. A comprehensive synonym guide (SYNONYM_GUIDE) listing clinical synonyms for all 46 symptoms was included in the system prompt. The model returned structured JSON with binary labels (1=present, 0=absent), with explicit instructions to exclude negated symptoms. Temperature was set to 0.0 to maximize output reproducibility.

*Gemini 3.5 Flash (LLM).* Gemini 3.5 Flash (gemini-3.5-flash, Google DeepMind [19]) was applied using the identical zero-shot protocol and SYNONYM_GUIDE as Claude Haiku. CC/HPI text was truncated to 2,500 characters per note (mean CC/HPI length: 1,847 characters; notes exceeding limit: n=312, 11.4%), with up to five retry attempts per note for transient API errors. Non-determinism at temperature=0.0 cannot be fully ruled out, as some LLM APIs retain stochastic elements in sampling; however, temperature=0.0 was used consistently to minimize output variance.

*Hybrid pipeline (LLM + Rule-based Negation Filtering).* As an exploratory investigation, a post-processing hybrid pipeline was developed to test whether rule-based negation filtering could further improve LLM precision beyond what contextual prompting already achieves. LLM predictions labeled as present (pred=1) were re-evaluated against the original CC/HPI text: if a matching symptom keyword was found within a 100-character window preceded by a negation cue (e.g., “denies,” “no,” “resolved”), the prediction was reversed to absent (pred=0). This was applied to both Claude Haiku and Gemini 3.5 Flash, yielding Claude+Hybrid and Gemini+Hybrid variants.

### Ground Truth Annotation

To construct the ground-truth dataset, 200 discharge notes were randomly sampled from the CRC cohort (random_state=42). CC/HPI sections were independently annotated by two expert annotators: a registered nurse with doctoral-level expertise in oncology symptom science (Y.L., PhD, RN) and a nursing professor (E.K., PhD, MPH, RN) with 9 years of clinical experience. A standardized coding manual was developed prior to annotation specifying: (1) symptom presence — any symptom explicitly mentioned as currently active in the CC/HPI; (2) symptom absence — any symptom explicitly denied, described as resolved, or not mentioned; and (3) negation rules — symptoms explicitly denied or resolved within the CC/HPI were coded absent, whereas constructions such as ‘no relief from nausea’ were coded present (the symptom is active despite the negation word). All 46 symptoms were labeled as binary values (1=present, 0=absent). Inter-rater reliability was assessed from these two sets of independent labels; all discordant labels were then resolved through joint adjudication of the source text by both annotators, and the resulting consensus labels constituted the ground-truth dataset. Inter-rater agreement was quantified using Cohen’s kappa, computed separately for each of the 46 symptoms based on the annotators’ independent, pre-adjudication labels. To assess the representativeness of this subsample, demographic and note-level characteristics of the 200 sampled notes were compared descriptively with those of the full analytic cohort.

### Reproducibility

To support reproducibility, all LLM-based extractions used fixed, versioned model identifiers (Claude Haiku: claude-haiku4.5; Gemini 3.5 Flash: gemini-3.5-flash) accessed via their respective vendor APIs [08/13/2026], with temperature fixed at 0.0 for both models to maximize output reproducibility. Gemini 3.5 Flash inputs were truncated to 2,500 characters per note, with up to five retry attempts per note for transient API errors. Ground-truth note sampling used a fixed random seed (random_state=42). All extraction pipelines, annotation guidelines, and analysis scripts are publicly available (see Code Availability).

### Evaluation

Performance was assessed using Macro F1, Micro F1, Macro Precision, and Macro Recall across all 46 symptoms against the 200 adjudicated gold-standard notes. Confusion matrices (TP, FP, FN, TN) were computed across all 200 notes (9,200 symptom–note pairs). Pairwise method comparisons used McNemar’s test on matched note-level binary predictions, with Bonferroni correction applied for multiple comparisons (adjusted α = 0.05/15 = 0.003).

### Ethical Considerations

Ethics statement: This study used the MIMIC-IV database under a valid data use agreement (DUA) with PhysioNet.[11] The study protocol was submitted to the University of Michigan Medicine Institutional Review Board, which determined that the study was not regulated. This determination reflects the study’s exclusive use of the publicly available, fully de-identified MIMIC-IV dataset, with no direct interaction with or identifiable information about human subjects. All data were securely stored and analyzed in a protected computing environment.

## Results

### Study Cohort

The final analytic sample comprised 2,704 discharge notes from 1,507 unique CRC patients (mean age 65.5 years [SD 15.0]; 51.4% male), following exclusion of 24 notes for which CC/HPI extraction failed due to non-standard formatting or absent section headers. Sample characteristics are presented in Table 2; further clinical and outcome characteristics relevant to the network and predictive-validity analyses are reported in a companion study.12 The 200-note gold-standard sample (147 unique patients) was broadly comparable to the full analytic cohort (1,507 patients) with respect to age (mean 64.9 [SD 15.3] vs. 65.5 [SD 15.0] years) and sex (48.3% vs. 51.4% male). Mean CC/HPI length in the 200-note sample was 982 characters (SD 640; range 13–3,774), somewhat shorter than the full-cohort mean reported above (1,847 characters); given the strongly right-skewed note-length distribution, this difference likely reflects a small number of unusually long notes in the full cohort rather than systematic undersampling of longer notes, but it is noted as a limitation of the representativeness check.

**Table 2.** Sample Characteristics of the Analytic Cohort (N=1,507 Patients, 2,704 Discharge Notes)

| Characteristic | N (%) or Mean (SD) |
| --- | --- |
| Patients, N | 1,507 |
| Discharge notes, N | 2,704 |
| Notes per patient, mean | 1.81 |
| Age, years, mean (SD) | 65.5 (15.0) |
| Sex |  |
| Male | 774 (51.4%) |
| Female | 733 (48.6%) |
| Race/ethnicity |  |
| White | 1,043 (69.2%) |
| Black/African American | 185 (12.3%) |
| Asian | 94 (6.2%) |
| Hispanic/Latino | 43 (2.9%) |
| Other/unknown/not reported | 142 (9.4%) |
| Insurance |  |
| Medicare | 723 (48.0%) |
| Private | 522 (34.6%) |
| Medicaid | 218 (14.5%) |
| Other/none/missing | 44 (2.9%) |
| Marital status |  |
| Married | 736 (48.8%) |
| Single | 449 (29.8%) |
| Widowed | 203 (13.5%) |
| **Divorced** | 91 (6.0%) |
| **Missing** | 28 (1.9%) |

*Note. Race/ethnicity, insurance, and marital status are as recorded in the MIMIC-IV hospital module at the patient's first CRC-related admission. Percentages are of the 1,507 unique patients and may not sum to 100% due to rounding or missing data.*

### Inter-Rater Reliability

Of 9,200 symptom-note pairs (200 notes × 46 symptoms) independently coded by both annotators, 277 (3.0%) were discordant. Pooled (micro-averaged) agreement across all symptom-note pairs was 97.0% (Cohen’s kappa = 0.71), indicating substantial overall agreement. However, the macro-averaged kappa (mean across the symptom-specific kappa values) was lower at 0.49, reflecting considerable variability in agreement across individual symptoms. Twenty of 46 symptoms (43%) showed substantial-to-almost-perfect agreement (kappa ≥ 0.60; e.g., cough kappa=0.94, vomiting kappa=0.92, nausea kappa=0.86), while 12 symptoms (26%) showed only slight agreement (kappa < 0.20) despite high raw percent agreement (94–99%), consistent with the known kappa paradox for low-prevalence binary ratings. Five symptoms (hair loss, altered self-image, problems with sexual interest/activity, erectile dysfunction, and nighttime urinary frequency) had zero positive instances from either annotator, precluding kappa estimation, and are reported as not estimable (kappa = NA); all 46 symptoms were retained in the analysis, with the macro-averaged kappa computed over the 41 symptoms with estimable values. All discordances were resolved via joint adjudication (see Supplementary Table2).

Symptom prevalence in the gold-standard sample was highly imbalanced. Consensus (post-adjudication) prevalence across the 46 symptoms ranged from 36.0% (abdominal pain) to 0% for six symptoms (difficulty concentrating, hair loss, altered self-image, problems with sexual interest/activity, erectile dysfunction, and nighttime urinary frequency); five of these six additionally had zero positive instances from either annotator prior to adjudication, precluding kappa estimation as noted above. This marked imbalance in symptom base rates underlies the divergence between macro-averaged (0.49) and pooled/micro-averaged (0.71) agreement reported above, and similarly explains why macro F1 (which weights all 46 symptoms equally) is consistently lower than micro F1 (which is dominated by the small number of frequently documented symptoms) for every extraction method reported below.

### Overall Symptom Extraction Performance

Table 3 presents overall performance across six extraction approaches. Figure 1 shows per-symptom F1 scores across all 46 symptoms for each method. Among the four original methods, Gemini 3.5 Flash achieved the highest Macro F1 (0.7035), Micro F1 (0.8591), and Macro Precision (0.7394), followed by Claude Haiku (Macro F1=0.6291, Macro Recall=0.7111). Both large language models substantially outperformed the traditional approaches, with rule-based (Macro F1=0.4427) and NER (0.3842) trailing well behind.

The hybrid variants yielded substantially lower performance than their LLM baselines — a negative finding that offers important methodological insight. Claude+Hybrid dropped from Macro F1=0.6291 to 0.5385, and Gemini+Hybrid from 0.7035 to 0.5752. Macro Recall fell sharply (Claude: 0.7111 → Claude+Hybrid: 0.5663; Gemini: 0.7105 → Gemini+Hybrid: 0.5287), driven by a large increase in false negatives. This pattern reveals a fundamental mismatch: the rule-based filter operates on a fixed 100-character window with a static negation cue list, which cannot reliably distinguish true negation from contextually positive mentions that incidentally contain negation words. For example, a note stating “no relief from nausea” conveys that nausea is present; the LLM correctly assigns pred=1, but the hybrid filter detects “no” within 100 characters of “nausea” and incorrectly flips the label to absent. Similarly, “patient denies any improvement in abdominal pain” is correctly identified as abdominal pain present by the LLM, but the rule-based window registers “denies” as a negation cue and overrides the prediction. These cases illustrate that the LLM’s contextual understanding of negation scope is superior to fixed-window pattern matching. Post-hoc rule-based negation filtering should not be applied to LLM outputs without syntactic scope validation, and this hybrid approach is not recommended for clinical NLP pipelines that use LLMs for symptom extraction.

**Table 3.** Overall Performance Comparison (GT: 200 Adjudicated Gold-Standard Notes, 46 Symptoms)

| Method | Macro F1 | Micro F1 | Macro Precision | Macro Recall |
| --- | --- | --- | --- | --- |
| Rule-based | 0.4427 | 0.6909 | 0.5062 | 0.4319 |
| NER (spaCy) | 0.3842 | 0.6031 | 0.4448 | 0.4061 |
| Claude Haiku | 0.6291 | 0.8121 | 0.6017 | 0.7111 |
| Claude + Hybrid | 0.5385 | 0.7149 | 0.5547 | 0.5663 |
| Gemini 3.5 Flash | 0.7035 | 0.8591 | 0.7394 | 0.7105 |
| Gemini + Hybrid | 0.5752 | 0.7590 | 0.6929 | 0.5287 |

*Note. Macro F1 averages F1 equally across 46 symptoms; Micro F1 weights by frequency. LLM methods received the symptom synonym guide as part of the prompt; rule-based and NER approaches used the same underlying symptom terminology through dictionary matching and post-processing, respectively. Hybrid variants apply post-hoc rule-based negation filtering to LLM predictions.*

**Figure 1.**
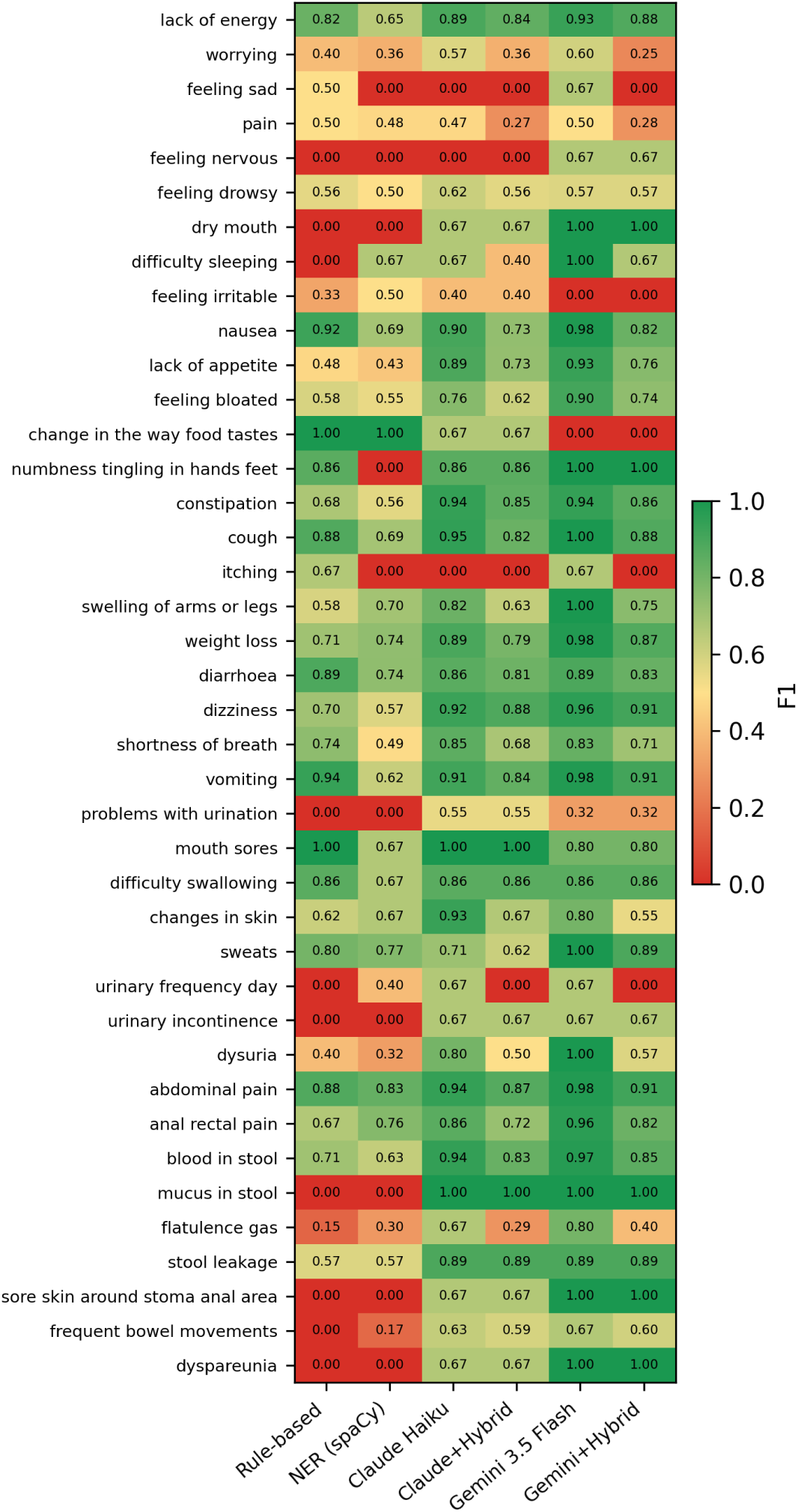
Per-Symptom F1 Score Heatmap by Method (GT: 200 Adjudicated Gold-Standard Notes) *Note: Each row represents one of 40 symptoms present in the gold standard; six zero-prevalence symptoms (F1 undefined, all methods trivially 0) are omitted for clarity but retained in the Table 3 macro-averages*.

### Confusion Matrix Analysis

Table 4 and Figure 2 present the confusion matrix across all 200 adjudicated gold-standard notes (9,200 pairs). The two LLMs produced the highest true-positive counts (Claude 495, Gemini 494), while rule-based and NER recovered fewer (351 and 332, respectively). NER generated the most false positives (221), whereas Gemini produced the fewest among the non-hybrid methods (108), giving it the highest precision. Hybrid negation filtering reduced false positives further (Gemini+Hybrid FP=75) but sharply increased false negatives (Claude+Hybrid FN=163 vs. Claude FN=53; Gemini+Hybrid FN=167 vs. Gemini FN=54), confirming the over-correction pattern.

**Table 4.**
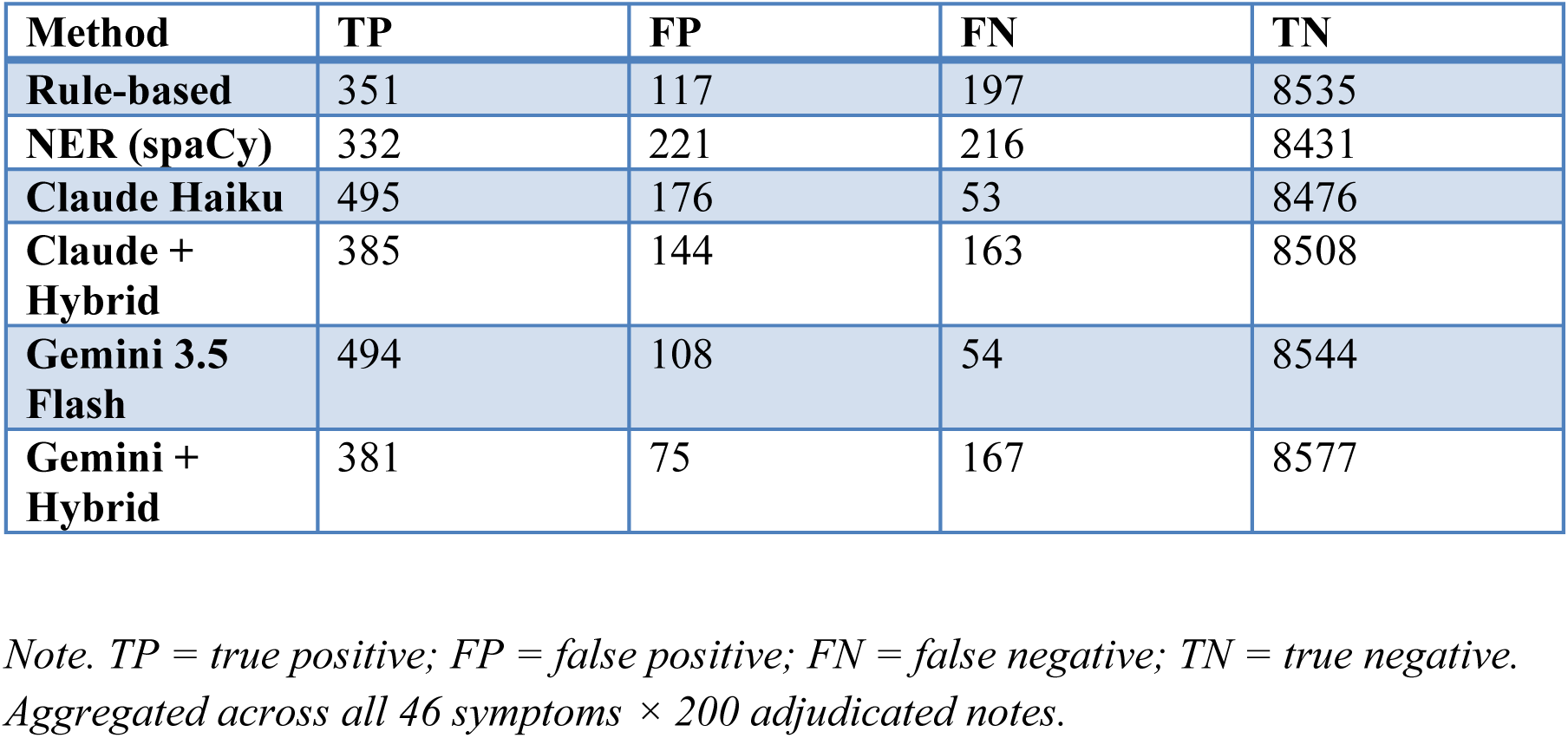
Confusion Matrix — GT 200 Adjudicated Notes (46 × 200 = 9,200 Symptom–Note Pairs)

| Method | TP | FP | FN | TN |
| --- | --- | --- | --- | --- |
| Rule-based | 351 | 117 | 197 | 8535 |
| NER (spaCy) | 332 | 221 | 216 | 8431 |
| Claude Haiku | 495 | 176 | 53 | 8476 |
| Claude + Hybrid | 385 | 144 | 163 | 8508 |
| Gemini 3.5 Flash | 494 | 108 | 54 | 8544 |
| Gemini + Hybrid | 381 | 75 | 167 | 8577 |

Note. TP = true positive; FP = false positive; FN = false negative; TN = true negative. Aggregated across all 46 symptoms × 200 adjudicated notes.

**Figure 2.**
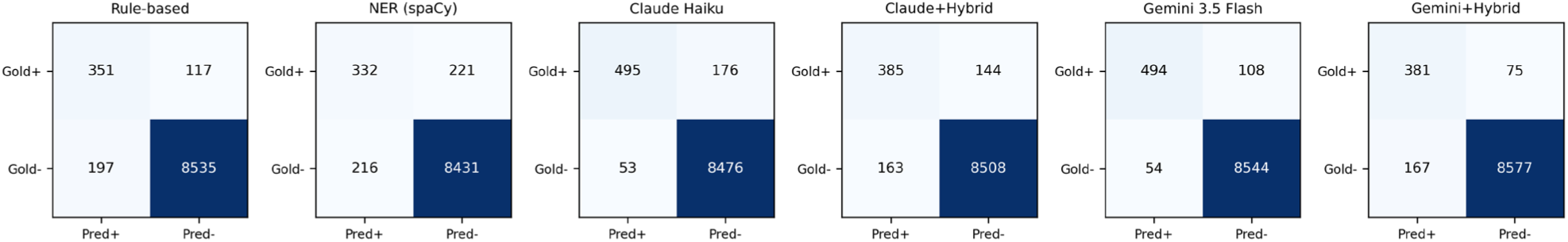
Confusion Matrix Heatmap. *Note. GT 200 Adjudicated Notes. Each panel shows the 2×2 confusion matrix for one method across all 46 symptoms × 200 notes (9,200 pairs). Darker blue = higher cell count*.

## Discussion

In this benchmarking study using 2,704 CRC discharge notes from MIMIC-IV, zero-shot LLM-based symptom extraction substantially outperformed traditional rule-based and NER approaches, with Gemini 3.5 Flash achieving the highest overall performance (Macro F1 = 0.70), followed by Claude Haiku (Macro F1 = 0.63). This finding is consistent with emerging evidence demonstrating the potential of LLMs for clinical information extraction from unstructured EHR narratives, including symptom extraction, with reduced reliance on task-specific model development.[20,21] Applying post-hoc rule-based negation filtering to LLM outputs (the hybrid approach) paradoxically degraded performance in our study, as fixed-window negation matching overrode some otherwise correct LLM predictions. This finding suggests that simple rule-based negation post-processing may not improve—and may even reduce—the performance of LLM-based symptom extraction when contextual information has already been incorporated into the model prediction. More sophisticated approaches that account for syntactic and contextual negation scope may therefore be needed when combining LLM extraction with rule-based post-processing.

Inter-rater reliability, assessed on the 200-note gold standard, was substantial when pooled across all symptoms (κ = 0.71) but showed wide symptom-level variability (macro-κ = 0.49), with several low-prevalence symptoms showing low κ despite high percent agreement. This discrepancy is consistent with the well-described prevalence effect on Cohen’s κ, whereby highly imbalanced category distributions can produce relatively low κ values despite high observed agreement.[22] The gold standard was constructed by two researchers who independently labeled symptom presence in each note according to a predefined annotation guideline, with discrepancies resolved through consensus adjudication. Establishing a reliable ground truth for clinical NLP requires clearly specified annotation guidelines and reproducible adjudication procedures, particularly when clinical concepts may be expressed implicitly or using variable terminology in narrative text.[23] Our findings further suggest that sufficient clinical domain expertise may be important for distinguishing such symptom expressions and applying symptom definitions consistently. Together, these findings underscore the importance of interpreting symptom-specific extraction performance alongside ground-truth reliability, particularly for rare symptoms.

Performance also varied considerably across individual symptoms (Figure 1), which the aggregate Macro and Micro F1 values in Table 3 do not fully capture. The general “pain” item is a clear example: despite reasonably high prevalence in the gold standard (18.0%), F1 for pain was only 0.47 for Claude Haiku and 0.50 for Gemini 3.5 Flash, well below the two anatomically site-specific pain items, abdominal_pain (F1 = 0.94 and 0.98, respectively) and anal_rectal_pain (F1 = 0.86 and 0.96, respectively), each of which exceeded 0.85 for both LLMs. Because the annotation guideline and both LLM system prompts specify that “pain” and the site-specific pain items are mutually exclusive—a mention of abdominal or anorectal pain alone should not also trigger the general pain label—this gap suggests that this category boundary is difficult to apply consistently in free-text narrative, where pain is not always clearly localized to a single anatomical site.

This difficulty was not unique to the LLMs: inter-rater agreement between the two human annotators was likewise substantially lower for pain (κ = 0.52, moderate) than for abdominal_pain (κ = 0.79, substantial) or anal_rectal_pain (κ = 0.86, almost perfect; Supplementary Table 2). The parallel pattern observed in both human annotation and LLM extraction suggests that the general-versus-site-specific distinction represents an inherently ambiguous aspect of the underlying symptom taxonomy rather than a failure specific to any one extraction method.

Macro F1 was also mechanically constrained by symptom prevalence. The six symptoms with zero positive instances in the 200-note gold standard necessarily yielded F1 = 0 for every extraction method because no method could register a true positive for a symptom that did not occur in the evaluation set. Because these six symptoms contributed equally to each method’s macro-averaged F1 in Table 3, the reported Macro F1 values for all four primary methods are lower than they would be if calculated only across adequately represented symptoms. This behavior reflects the class-wise averaging inherent to Macro F1, in which performance for each evaluated class contributes equally to the aggregate score regardless of class frequency.[24] Thus, the lower aggregate Macro F1 partly reflects the composition and size of the available 200-note gold standard rather than the performance of any individual extraction method alone. These findings underscore the value of larger evaluation sets with sufficient representation of rare symptoms when benchmarking broad symptom inventories.

### Limitations

This study has several limitations. First, although LLMs were applied in a zero-shot setting to maximize generalizability, fine-tuned clinical models may achieve higher extraction accuracy. Second, the large language models benchmarked here are proprietary, versioned, and subject to deprecation: Gemini 2.5 Flash was retired by the provider during this study and superseded by Gemini 3.5 Flash, and cloud-based APIs also require transmitting clinical text to external servers. Such version churn poses reproducibility and data-governance challenges for LLM-based clinical pipelines; we mitigate this by reporting exact model versions and releasing our prompts and extraction code, though locally deployable open-weight models may ultimately offer more durable and privacy-preserving alternatives. Finally, this study was conducted at a single academic medical center (BIDMC), and generalizability to community or international settings requires validation. Fourth, although all methods were mapped to the same predefined 46-symptom inventory, the manner in which symptom terminology was supplied differed across approaches -- LLM methods received the synonym guide directly in the prompt, whereas rule-based and NER methods used the same terminology through dictionary matching and post-hoc mapping, respectively -- reflecting the distinct architectures of these approaches and potentially influencing comparative performance.

### Implications for Practice

The application of LLM-based symptom extraction to EHR data could enable longitudinal monitoring of symptom occurrence, prevalence, and trajectories across the cancer care continuum using routinely collected clinical notes. Linking these symptom patterns with patients’ demographic, clinical, treatment, and social characteristics could help identify individuals experiencing persistent, worsening, or disproportionate symptom burden who may be at greater risk for adverse outcomes. This approach could provide a foundation for the early identification of high-risk patients and inform timely, tailored symptom management interventions based on individual symptom trajectories and care needs. Ultimately, integrating LLM-based symptom surveillance into EHR systems may support more proactive, personalized, and equitable symptom management in oncology practice.

## Ethics and Data Availability

Ethics statement: This study used the MIMIC-IV database under a valid data use agreement (DUA) with PhysioNet.[11] The study protocol was submitted to the University of Michigan Medicine Institutional Review Board, which determined that the study was not regulated. This determination reflects the study’s exclusive use of the publicly available, fully de-identified MIMIC-IV dataset, with no direct interaction with or identifiable information about human subjects.

## Data availability

The MIMIC-IV dataset is publicly available at https://physionet.org/content/mimiciv/ to credentialed researchers who complete the required training and data use agreement.

## Code availability

All extraction pipelines, annotation guidelines, and analysis scripts are publicly available at https://github.com/youranrl-dot/crc_symptom_llm.

## Author contributions

Y.L.: Conceptualization, Methodology, Software, Formal analysis, Writing — original draft, Writing — review and editing. I.D.: Methodology, Writing — review and editing. X.H.: Methodology, Formal analysis. Y.J.: Supervision, Writing — review and editing.

## Acknowledgement

We would like to thank Eungyung Kim, for her valuable contribution to the manual annotation of the 200 clinical notes used to construct the ground-truth dataset.

---

**Supplementary Table 1.**
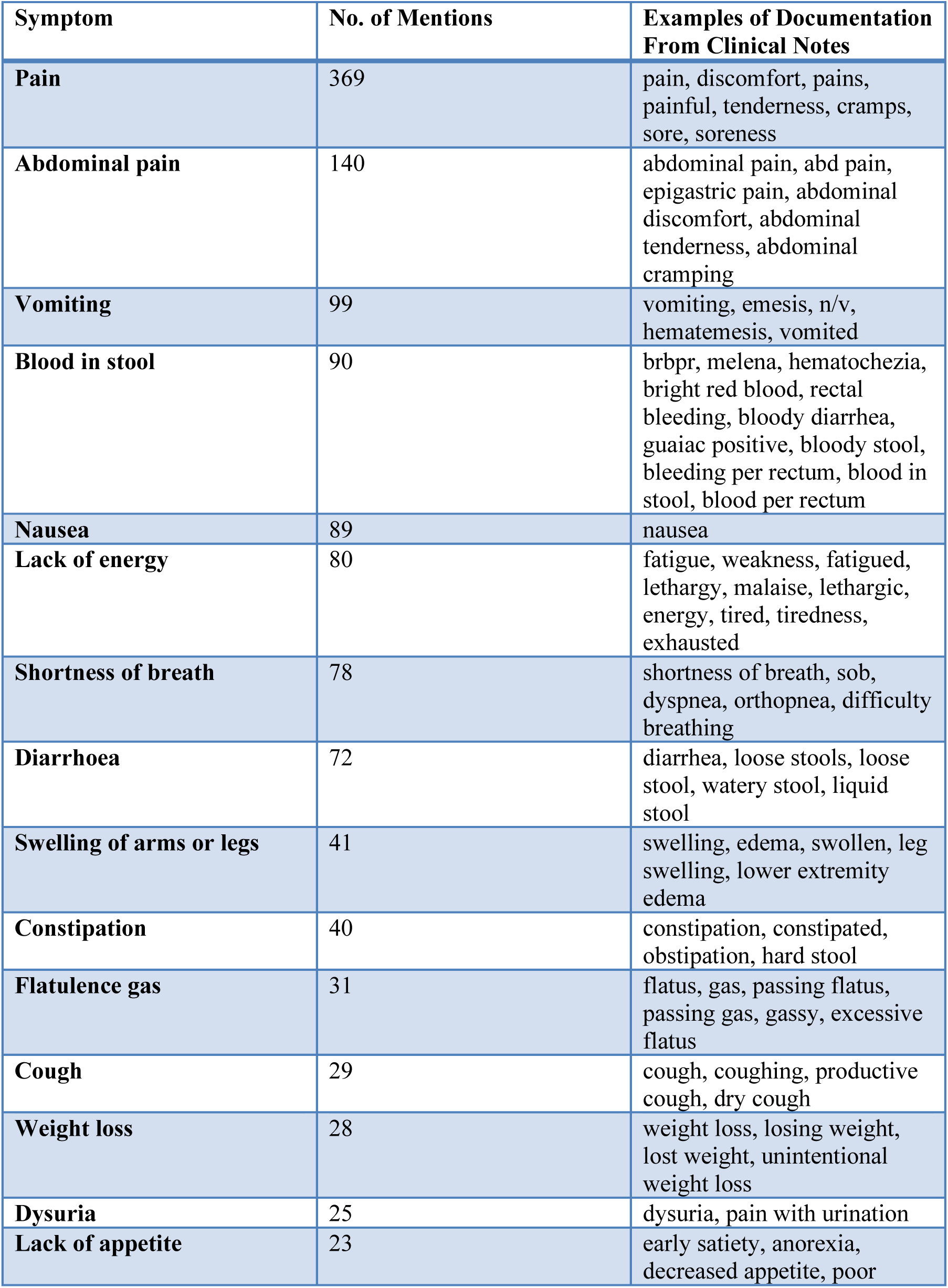
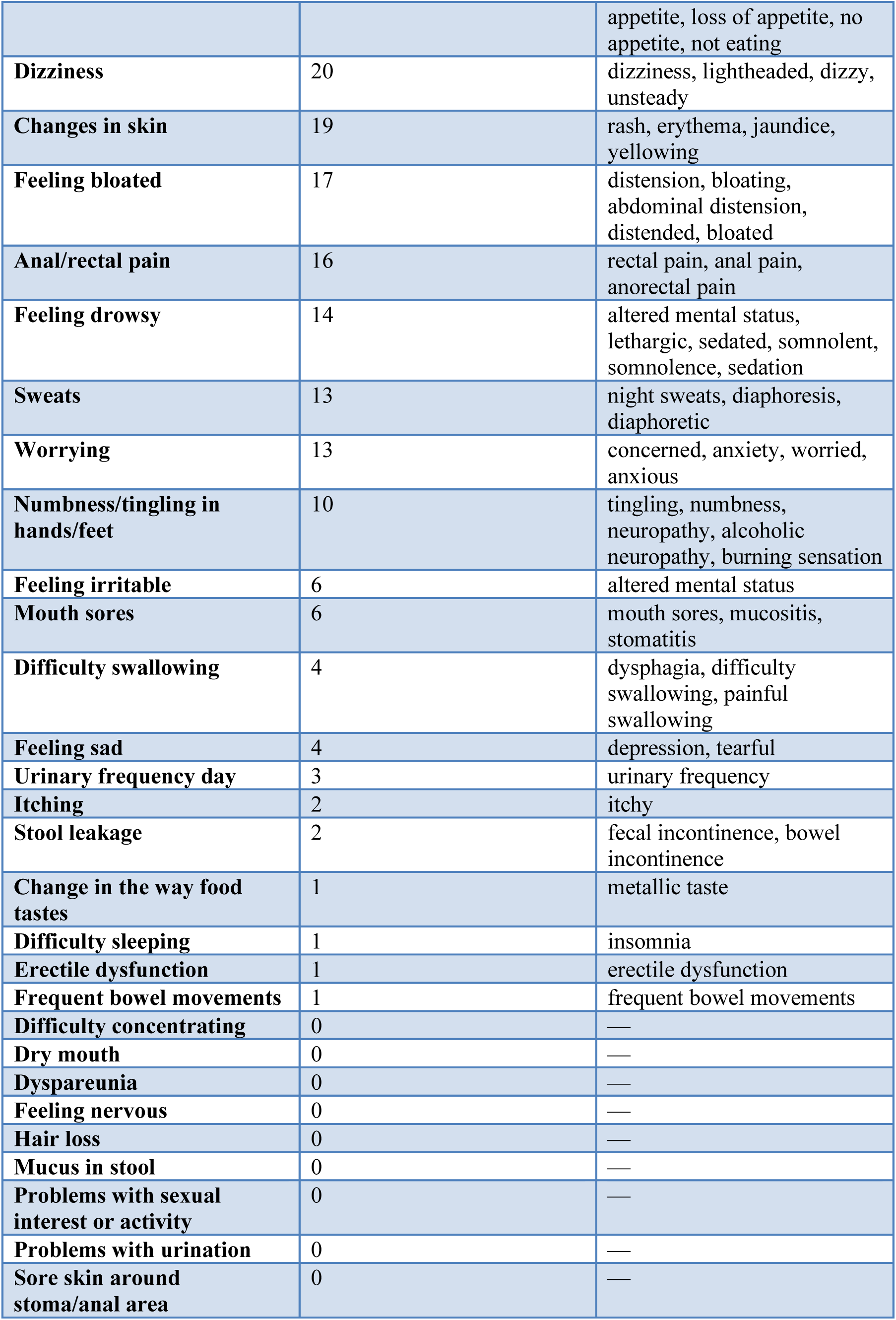
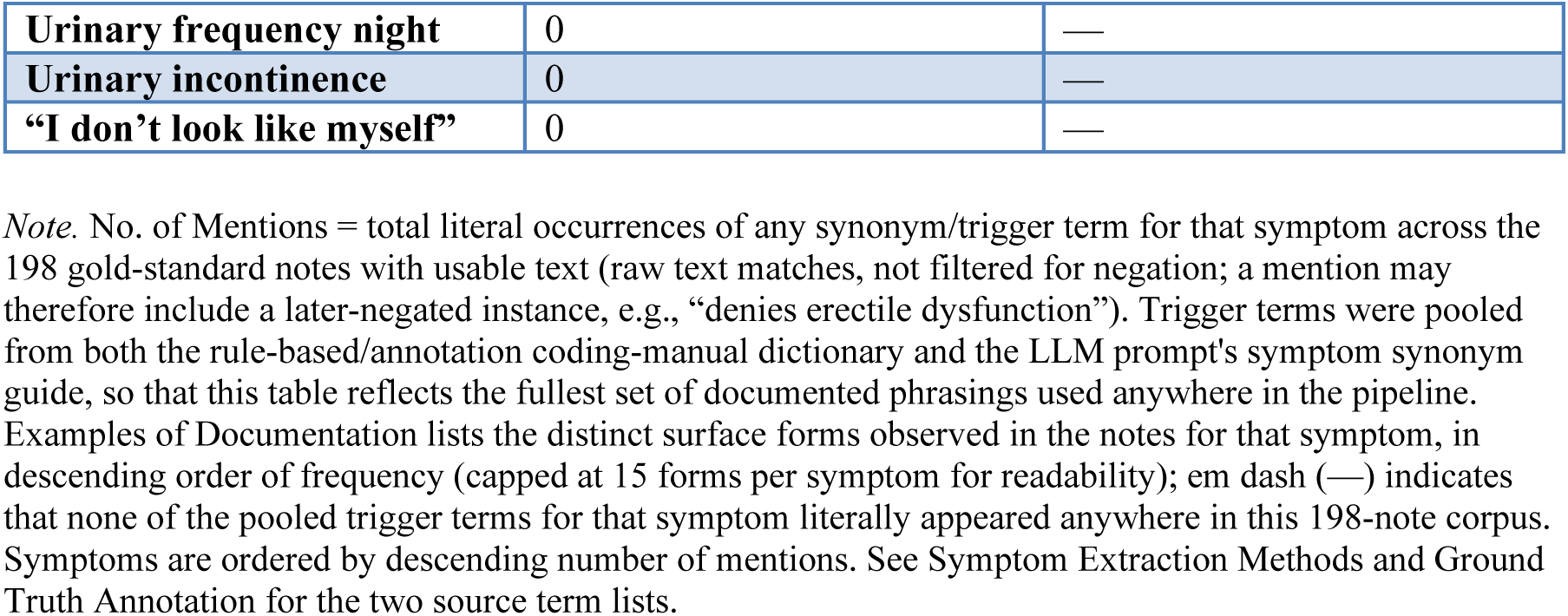
Symptom Documentation Examples From the 200-Note Gold-Standard Corpus.

**Supplementary Table 2.**
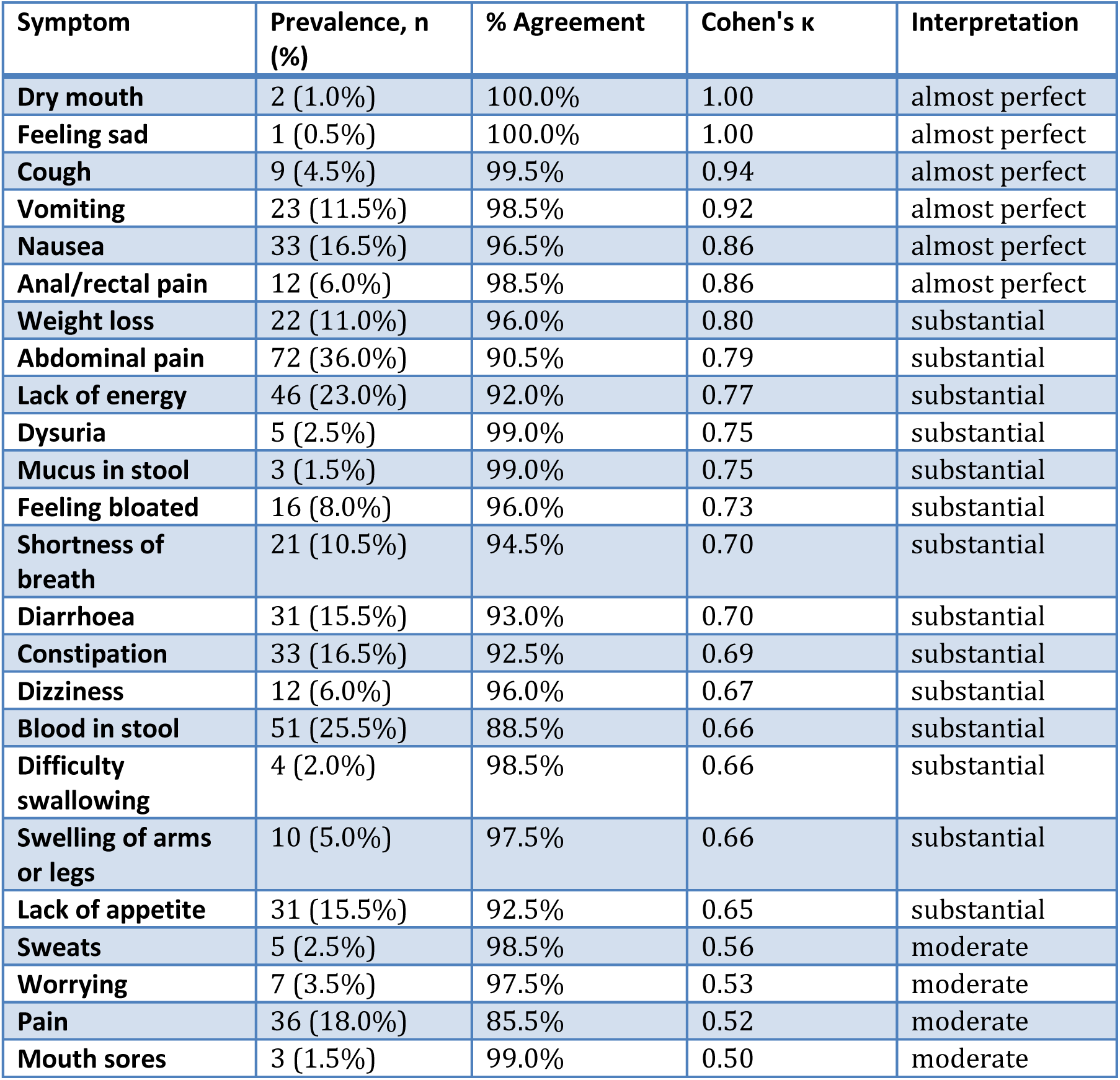
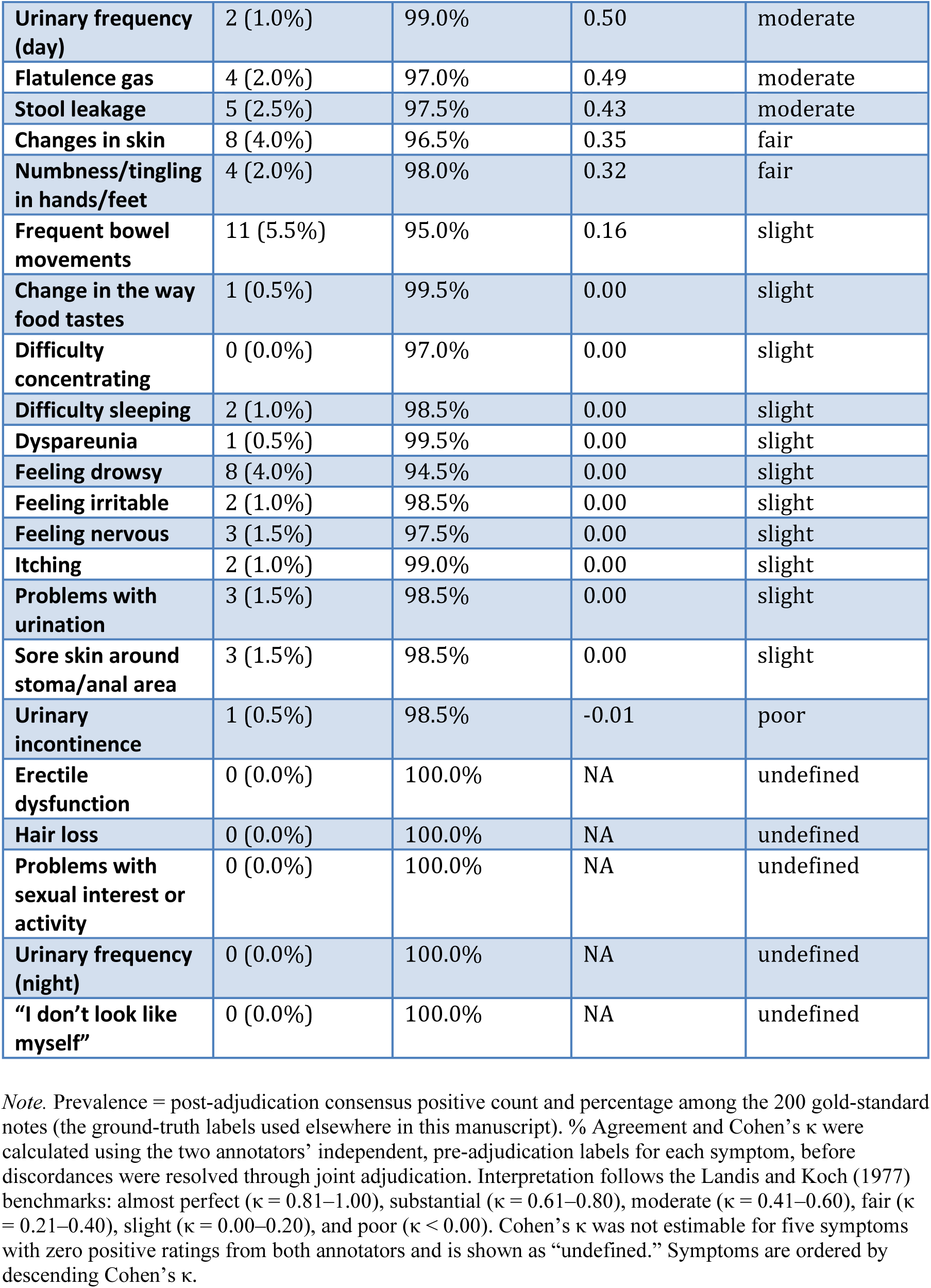
Per-Symptom Inter-Rater Reliability, Percent Agreement, and Prevalence (200-Note Gold Standard)

