## Supplemental Table for "Leveraging Large Language Models for Colorectal Cancer Symptom Extraction from MIMIC-IV Clinical Notes"

**Supplementary Table 1. Symptom Documentation Examples From the 200-Note Gold-Standard Corpus**

| Symptom | No. of Mentions | Examples of Documentation From Clinical Notes |
|---|---|---|
| **Pain** | 369 | pain, discomfort, pains, painful, tenderness, cramps, sore, soreness |
| **Abdominal pain** | 140 | abdominal pain, abd pain, epigastric pain, abdominal discomfort, abdominal tenderness, abdominal cramping |
| **Vomiting** | 99 | vomiting, emesis, n/v, hematemesis, vomited |
| **Blood in stool** | 90 | brbpr, melena, hematochezia, bright red blood, rectal bleeding, bloody diarrhea, guaiac positive, bloody stool, bleeding per rectum, blood in stool, blood per rectum |
| **Nausea** | 89 | nausea |
| **Lack of energy** | 80 | fatigue, weakness, fatigued, lethargy, malaise, lethargic, energy, tired, tiredness, exhausted |
| **Shortness of breath** | 78 | shortness of breath, sob, dyspnea, orthopnea, difficulty breathing |
| **Diarrhoea** | 72 | diarrhea, loose stools, loose stool, watery stool, liquid stool |
| **Swelling of arms or legs** | 41 | swelling, edema, swollen, leg swelling, lower extremity edema |
| **Constipation** | 40 | constipation, constipated, obstipation, hard stool |
| **Flatulence gas** | 31 | flatus, gas, passing flatus, passing gas, gassy, excessive flatus |
| **Cough** | 29 | cough, coughing, productive cough, dry cough |
| **Weight loss** | 28 | weight loss, losing weight, lost weight, unintentional weight loss |
| **Dysuria** | 25 | dysuria, pain with urination |
| **Lack of appetite** | 23 | early satiety, anorexia, decreased appetite, poor |

| | | |
|---|---|---|
| | | appetite, loss of appetite, no appetite, not eating |
| **Dizziness** | 20 | dizziness, lightheaded, dizzy, unsteady |
| **Changes in skin** | 19 | rash, erythema, jaundice, yellowing |
| **Feeling bloated** | 17 | distension, bloating, abdominal distension, distended, bloated |
| **Anal/rectal pain** | 16 | rectal pain, anal pain, anorectal pain |
| **Feeling drowsy** | 14 | altered mental status, lethargic, sedated, somnolent, somnolence, sedation |
| **Sweats** | 13 | night sweats, diaphoresis, diaphoretic |
| **Worrying** | 13 | concerned, anxiety, worried, anxious |
| **Numbness/tingling in hands/feet** | 10 | tingling, numbness, neuropathy, alcoholic neuropathy, burning sensation |
| **Feeling irritable** | 6 | altered mental status |
| **Mouth sores** | 6 | mouth sores, mucositis, stomatitis |
| **Difficulty swallowing** | 4 | dysphagia, difficulty swallowing, painful swallowing |
| **Feeling sad** | 4 | depression, tearful |
| **Urinary frequency day** | 3 | urinary frequency |
| **Itching** | 2 | itchy |
| **Stool leakage** | 2 | fecal incontinence, bowel incontinence |
| **Change in the way food tastes** | 1 | metallic taste |
| **Difficulty sleeping** | 1 | insomnia |
| **Erectile dysfunction** | 1 | erectile dysfunction |
| **Frequent bowel movements** | 1 | frequent bowel movements |
| **Difficulty concentrating** | 0 | — |
| **Dry mouth** | 0 | — |
| **Dyspareunia** | 0 | — |
| **Feeling nervous** | 0 | — |
| **Hair loss** | 0 | — |
| **Mucus in stool** | 0 | — |
| **Problems with sexual interest or activity** | 0 | — |
| **Problems with urination** | 0 | — |
| **Sore skin around stoma/anal area** | 0 | — |

| | | |
|---|---|---|
| **Urinary frequency night** | 0 | — |
| **Urinary incontinence** | 0 | — |
| **"I don't look like myself"** | 0 | — |

*Note.* No. of Mentions = total literal occurrences of any synonym/trigger term for that symptom across the 198 gold-standard notes with usable text (raw text matches, not filtered for negation; a mention may therefore include a later-negated instance, e.g., "denies erectile dysfunction"). Trigger terms were pooled from both the rule-based/annotation coding-manual dictionary and the LLM prompt's symptom synonym guide, so that this table reflects the fullest set of documented phrasings used anywhere in the pipeline. Examples of Documentation lists the distinct surface forms observed in the notes for that symptom, in descending order of frequency (capped at 15 forms per symptom for readability); em dash (—) indicates that none of the pooled trigger terms for that symptom literally appeared anywhere in this 198-note corpus. Symptoms are ordered by descending number of mentions. See Symptom Extraction Methods and Ground Truth Annotation for the two source term lists.

**Supplementary Table 2. Per-Symptom Inter-Rater Reliability, Percent Agreement, and Prevalence (200-Note Gold Standard)**

| Symptom | Prevalence, n (%) | % Agreement | Cohen's κ | Interpretation |
|---|---|---|---|---|
| **Dry mouth** | 2 (1.0%) | 100.0% | 1.00 | almost perfect |
| **Feeling sad** | 1 (0.5%) | 100.0% | 1.00 | almost perfect |
| **Cough** | 9 (4.5%) | 99.5% | 0.94 | almost perfect |
| **Vomiting** | 23 (11.5%) | 98.5% | 0.92 | almost perfect |
| **Nausea** | 33 (16.5%) | 96.5% | 0.86 | almost perfect |
| **Anal/rectal pain** | 12 (6.0%) | 98.5% | 0.86 | almost perfect |
| **Weight loss** | 22 (11.0%) | 96.0% | 0.80 | substantial |
| **Abdominal pain** | 72 (36.0%) | 90.5% | 0.79 | substantial |
| **Lack of energy** | 46 (23.0%) | 92.0% | 0.77 | substantial |
| **Dysuria** | 5 (2.5%) | 99.0% | 0.75 | substantial |
| **Mucus in stool** | 3 (1.5%) | 99.0% | 0.75 | substantial |
| **Feeling bloated** | 16 (8.0%) | 96.0% | 0.73 | substantial |
| **Shortness of breath** | 21 (10.5%) | 94.5% | 0.70 | substantial |
| **Diarrhoea** | 31 (15.5%) | 93.0% | 0.70 | substantial |
| **Constipation** | 33 (16.5%) | 92.5% | 0.69 | substantial |
| **Dizziness** | 12 (6.0%) | 96.0% | 0.67 | substantial |
| **Blood in stool** | 51 (25.5%) | 88.5% | 0.66 | substantial |
| **Difficulty swallowing** | 4 (2.0%) | 98.5% | 0.66 | substantial |
| **Swelling of arms or legs** | 10 (5.0%) | 97.5% | 0.66 | substantial |
| **Lack of appetite** | 31 (15.5%) | 92.5% | 0.65 | substantial |
| **Sweats** | 5 (2.5%) | 98.5% | 0.56 | moderate |
| **Worrying** | 7 (3.5%) | 97.5% | 0.53 | moderate |
| **Pain** | 36 (18.0%) | 85.5% | 0.52 | moderate |
| **Mouth sores** | 3 (1.5%) | 99.0% | 0.50 | moderate |

| | | | | |
|---|---|---|---|---|
| **Urinary frequency (day)** | 2 (1.0%) | 99.0% | 0.50 | moderate |
| **Flatulence gas** | 4 (2.0%) | 97.0% | 0.49 | moderate |
| **Stool leakage** | 5 (2.5%) | 97.5% | 0.43 | moderate |
| **Changes in skin** | 8 (4.0%) | 96.5% | 0.35 | fair |
| **Numbness/tingling in hands/feet** | 4 (2.0%) | 98.0% | 0.32 | fair |
| **Frequent bowel movements** | 11 (5.5%) | 95.0% | 0.16 | slight |
| **Change in the way food tastes** | 1 (0.5%) | 99.5% | 0.00 | slight |
| **Difficulty concentrating** | 0 (0.0%) | 97.0% | 0.00 | slight |
| **Difficulty sleeping** | 2 (1.0%) | 98.5% | 0.00 | slight |
| **Dyspareunia** | 1 (0.5%) | 99.5% | 0.00 | slight |
| **Feeling drowsy** | 8 (4.0%) | 94.5% | 0.00 | slight |
| **Feeling irritable** | 2 (1.0%) | 98.5% | 0.00 | slight |
| **Feeling nervous** | 3 (1.5%) | 97.5% | 0.00 | slight |
| **Itching** | 2 (1.0%) | 99.0% | 0.00 | slight |
| **Problems with urination** | 3 (1.5%) | 98.5% | 0.00 | slight |
| **Sore skin around stoma/anal area** | 3 (1.5%) | 98.5% | 0.00 | slight |
| **Urinary incontinence** | 1 (0.5%) | 98.5% | -0.01 | poor |
| **Erectile dysfunction** | 0 (0.0%) | 100.0% | NA | undefined |
| **Hair loss** | 0 (0.0%) | 100.0% | NA | undefined |
| **Problems with sexual interest or activity** | 0 (0.0%) | 100.0% | NA | undefined |
| **Urinary frequency (night)** | 0 (0.0%) | 100.0% | NA | undefined |
| **"I don't look like myself"** | 0 (0.0%) | 100.0% | NA | undefined |

*Note.* Prevalence = post-adjudication consensus positive count and percentage among the 200 gold-standard notes (the ground-truth labels used elsewhere in this manuscript). % Agreement and Cohen's κ were calculated using the two annotators' independent, pre-adjudication labels for each symptom, before discordances were resolved through joint adjudication. Interpretation follows the Landis and Koch (1977) benchmarks: almost perfect (κ = 0.81–1.00), substantial (κ = 0.61–0.80), moderate (κ = 0.41–0.60), fair (κ = 0.21–0.40), slight (κ = 0.00–0.20), and poor (κ < 0.00). Cohen's κ was not estimable for five symptoms with zero positive ratings from both annotators and is shown as "undefined." Symptoms are ordered by descending Cohen's κ.
